# Modelling to support decisions about the geographic and demographic extensions of seasonal malaria chemoprevention in Benin

**DOI:** 10.1101/2024.04.25.24306333

**Authors:** Jeanne Lemant, Clara Champagne, William Houndjo, Julien Aïssan, Rock Aïkpon, Camille Houetohossou, Sakariahou Kpanou, Roland Goers, Cyriaque Affoukou, Emilie Pothin

## Abstract

**Background:** Seasonal malaria chemoprevention (SMC) has been implemented yearly in northern Benin since 2019 to reduce the malaria burden in children under 5 years of age. Its geographic scope was progressively extended until in 2022 two different extensions of SMC were considered: either demographic - children aged 5 to 10 in the currently targeted departments would also receive SMC, or geographic to children under 5 in new eligible departments to the south. As SMC had neither been implemented in the areas nor age groups suggested for expansion, modelling was used to compare the likely impact of both extensions.

**Methods:** The model OpenMalaria was calibrated to represent the history of malaria interventions and transmission risk in Benin. Currently planned future interventions and two scenarios for SMC extensions were simulated to inform where impact would be the highest.

**Results:** The model predicted that between 2024 and 2026 the geographic extension of SMC would avert at least four times more severe malaria cases and five times more direct malaria deaths per targeted child than the demographic extension. However, numbers of severe cases averted per targeted child were similar between health zones eligible for geographic extension.

**Conclusions:** The geographic extension is more impactful and likely more cost-effective than the demographic extension, and will be implemented from 2024. Health zones were prioritised by availability of community health workers to deliver SMC. Mathematical modelling was a supportive tool to understand the relative impact of the different proposed SMC extensions and contributed to the decision-making process. Its integration significantly enhanced the utilisation of data for decision-making purposes. Rather than being used for forecasting, the model provided qualitative guidance that complemented other types of evidence.

## Introduction

Malaria killed 608 000 persons in 2022, of which 76% were children under 5 [1]. This parasitic disease is one of the leading cause of death in children under 5 [2]. Strong reductions in malaria burden have been achieved since 2000, mainly thanks to insecticide-treated nets (ITNs) and artemisinin-based combinations therapies [3], but since 2015 progress has stalled. Insecticide and drug resistance, invasion of new vector species, and lack of funding threaten to reverse the decreasing trend. Additional interventions to nets and treatments are now required to further decrease transmission and protect those most at risk of severe malaria. Seasonal malaria chemoprevention (SMC) consists of administering antimalarials to all children living in seasonal transmission areas during the period of heightened malaria risk to clear existing infections and prevent new ones [4]. The number of children treated increased from 0.2 million in 2012 when the World Health Organisation (WHO) recommended SMC to 49 million ten years later. In Benin, where malaria is the first cause of consultation and which accounts for 2.1% of global malaria cases and 1.8% of deaths [1], SMC was first implemented in 2019 in the health zones of Malanville-Karimama (Alibori) and Tanguieta-Materi-Cobly (Atacora). The geographic scope of SMC was progressively extended until in 2021 almost 600 000 children under 5 living in the two Sahelian departments of Alibori and Atacora were targeted (NMCP, personal communication).

In 2022 the WHO updated their recommendations for SMC: the intervention is no longer recommended only to children under 5 years of age, but to all children at high risk of severe malaria. This increased flexibility is an opportunity for malaria endemic countries to better tailor their strategy to local needs. However, these new recommendations also mean National Malaria Control Programmes (NMCPs) need to make more informed choices. Three months after the release of the updated recommendations, the Benin NCMP initiated a collaboration with the Swiss Tropical and Public Health Institute to analyse together the relative benefits of either extending SMC to children under 10 years of age in Alibori and Atacora, where SMC was already implemented, or extending SMC to new eligible zones in departments of Borgou, Collines, and Donga but only targeting children under 5. Choosing between these the two options was necessary as the entire population of Benin is at risk for malaria, but funding is limited. This limited funding also constrained the number of new zones which could be targeted, so the NMCP was interested in a ranking of the new eligible zones based on impact to inform on the zone prioritisation.

SMC has been established as an efficacious intervention in numerous randomised controlled trials in children under 5 and under 10. Cissé et al. [5] administered SMC to children under 10 and observed incidence in children under 5 decrease by 57% and in children between 5 and 9 by 61% during the transmission season, while others found even larger reductions [6-8]. These differences in impact may be due to different settings and contexts. SMC is also an operationally feasible intervention which can reach coverages above 90% [9, 10].

To our knowledge, no study has compared the benefits of extending the age limit of SMC to expanding to new zones, but NMCPs need to know which strategy is more effective in their context to make an informed decision. While young children are usually found to be at higher risk for severe malaria [11, 12] the impact of SMC in different age groups may be influenced by factors such as intensity of transmission, seasonality or concomitant deployment of other interventions. In this context, all relevant factors must be assessed before targeting a specific age group. Beyond determining which of the two extension strategies would save more lives, understanding their relative impact would enable the NMCPs to weigh this likely impact by the cost of the intervention and choose the most cost-effective strategy. Such a quantification will intrinsically be context-specific and as such requires a method which can be adapted to the setting of interest. Mathematical modelling can mimic the past and current malaria burden in administrative units of a country and simulate the impact of various interventions, and has already been used to support decision-making [13, 14].

Our goal was to determine which extension of SMC would have more impact and be more cost-effective in Benin, and to rank new eligible zones according to the impact SMC could have.

## Methods

We used OpenMalaria to simulate the impact of interventions on malaria burden. OpenMalaria comprises an agent-based model of human malaria infections using a within-host component [15], together with a compartmental model to govern the evolution of the *Anopheles* population [16]. Simulations were set up and run using the R package openMalariaUtilities. The model was adapted to Benin at the commune level to make predictions tailored to the context of the country. The entire workflow is summarised in Figure 1.

**Figure 1:**
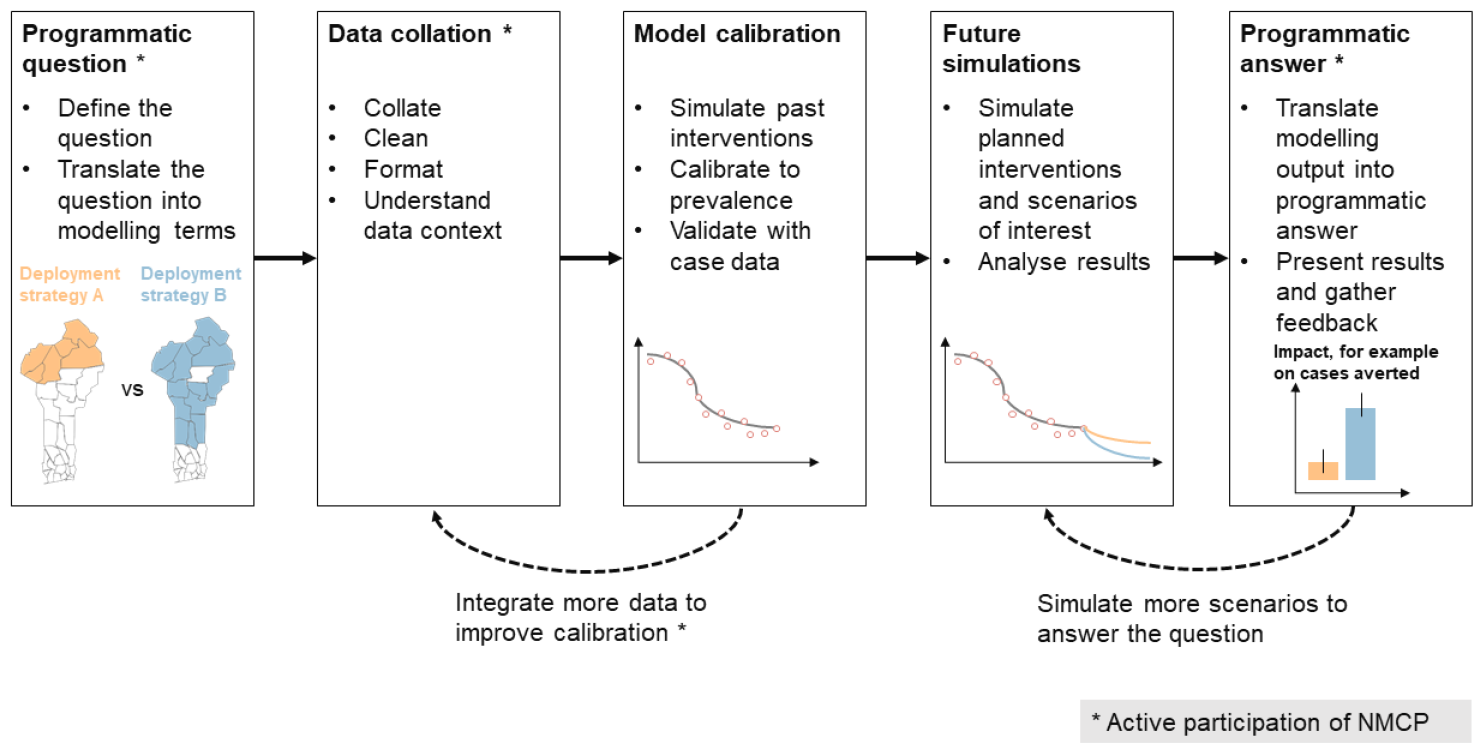
Applied workflow to answer programmatic questions with country-specific mathematical modelling. Schematics are for illustrative purpose only.

### Model parameterisation: capturing historical trends for each commune up to 2023

We collated global data on interventions efficacy and local data on past deployments in Benin and adjusted the modelled transmission risk in each commune so that yearly modelled prevalence would match estimates. These prevalence estimates in children aged 2 to 10 were post-processed by the Malaria Atlas Project from prevalence surveys and covariates [17] for years 2006 to 2019.

Dominant vector species in Benin are *Anopheles gambiae sensu lato and Anopheles funestus* [18]; in the model they were considered to contribute at 90% and 10% to transmission respectively. To account crudely for variation in mosquito behaviour, 10% of mosquito bites were assumed to take place outdoors. Intense pyrethroid resistance was first recorded by sentinel sites in 2016 (NMCP, personal communication) so from this year on vectors were assumed to be fully resistant to pyrethroids, with nets acting only as a physical barrier. Before that, we assumed vectors were moderately resistant to pyrethroids, using a previously published parameterisation of ITNs [19].

Seasonality of transmission was informed by reported incidence in each department, with a 15-day lag to account for the time between infection and onset of symptoms.

We used ITN use estimates from the Malaria Atlas Project [20] at the department level until the first mass distribution campaign in 2011. We then relied on household surveys [21, 22] to estimate ITN use, assuming nets distributed during each mass campaign had a three-year half-life. From 2020 not only ITNs were distributed but also piperonyl butoxide (PBO), and Interceptor G2 in 2023. Both these next-generation nets were modelled as restoring most susceptibility in vectors, based on an experimental hut trial recording 70% mortality in bioassays [19]. Durability based on net fabric was also taken into account, with nets made of polyethylene or polyester having a half-life of two - respectively 1.6 – years [23]. In 2023 most distributed nets were made of polyethylene except for one brand of PBO nets distributed in some northern communes. We made the conservative assumption that use of nets had remained constant since the last available survey in 2017. IRS active ingredient, population coverage and month of spraying were modelled at the commune level until the intervention was discontinued in 2022 [24]. Insecticide efficacy parameterisations were derived from experimental hut trials results [25].

Access to treatment in case of fever has been reported in household surveys between 2001 and 2017 [21, 22] at the department level. Once an individual seeks treatment, they do not necessarily receive a completely curative treatment due to care-seeking behaviour in unofficial structures, lack of compliance of care providers, lack of adherence of certain patients, presence of counterfeit medicines and imperfect efficacy of treatments. The effective treatment coverage used in the model is therefore the product of the access to health services measured during each of the five surveys by those factors which quantify the inefficiencies in the care management cascade [26]. After 2017 effective treatment coverage was assumed to remain constant. While access to treatment for uncomplicated malaria cases was derived from household surveys, for severe malaria cases we assumed that 48% of patients seek care [27].

After Malanville-Karimama (Alibori) and Tanguieta-Materi-Cobly (Atacora) in 2019, SMC was extended to Banikoara and Kandi-Gogounou-Segbana (Alibori) in 2020, and finally all children under 5 of both departments were targeted from 2021. Efficacy of SMC was modelled through a blood-clearance effect lasting 25 days to reproduce the effect size observed in a trial in the Sahel [28]. This means SMC is assumed to clear all infections for 25 days, then its effect wanes instantly. To match reported coverage data from the 2021 SMC campaign, 80% of eligible children under 5 were randomly selected to receive SMC at each of the four rounds from July to October.

The model reproduces the population age structure from the Benin 2013 census [29]. We also assumed 10 infections per 1000 people per year were imported in each commune.

The transmission level, represented by the pre-intervention Entomological Inoculation Rate (EIR) in 2000, was then adjusted for each commune to match the time series of past prevalence estimates. We also estimated lower and higher bounds for transmission intensity to represent the uncertainty in prevalence estimates and stochasticity of the model.

### Simulation of SMC extensions

Additionally to already planned interventions (mass net distribution campaign in 2026 and SMC administered to children under 5 in Alibori and Atacora, Table 1), we simulated both extension scenarios of SMC starting in 2024. The administration period, coverage, and efficacy were assumed to be identical to the already planned SMC campaigns. For the demographic extension of SMC we simulated its administration to children under 10 in Alibori and Atacora. For the geographic extension the NMCP had already identified the eligible zones situated in the region of the Sahel and where more than half the cases were occurring within four consecutive months. We simulated SMC administered to children under 5 in Borgou, Collines, and Donga except in the sanitary zone of Bembereke-Sinende (Borgou). This zone was already part of the Plus Project, a pilot study of perennial malaria chemoprevention (PMC) which should last until 2025 and targets children until 2 years, so we excluded this zone from the geographic extension as SMC and PMC are mutually exclusive.

**Table 1:**
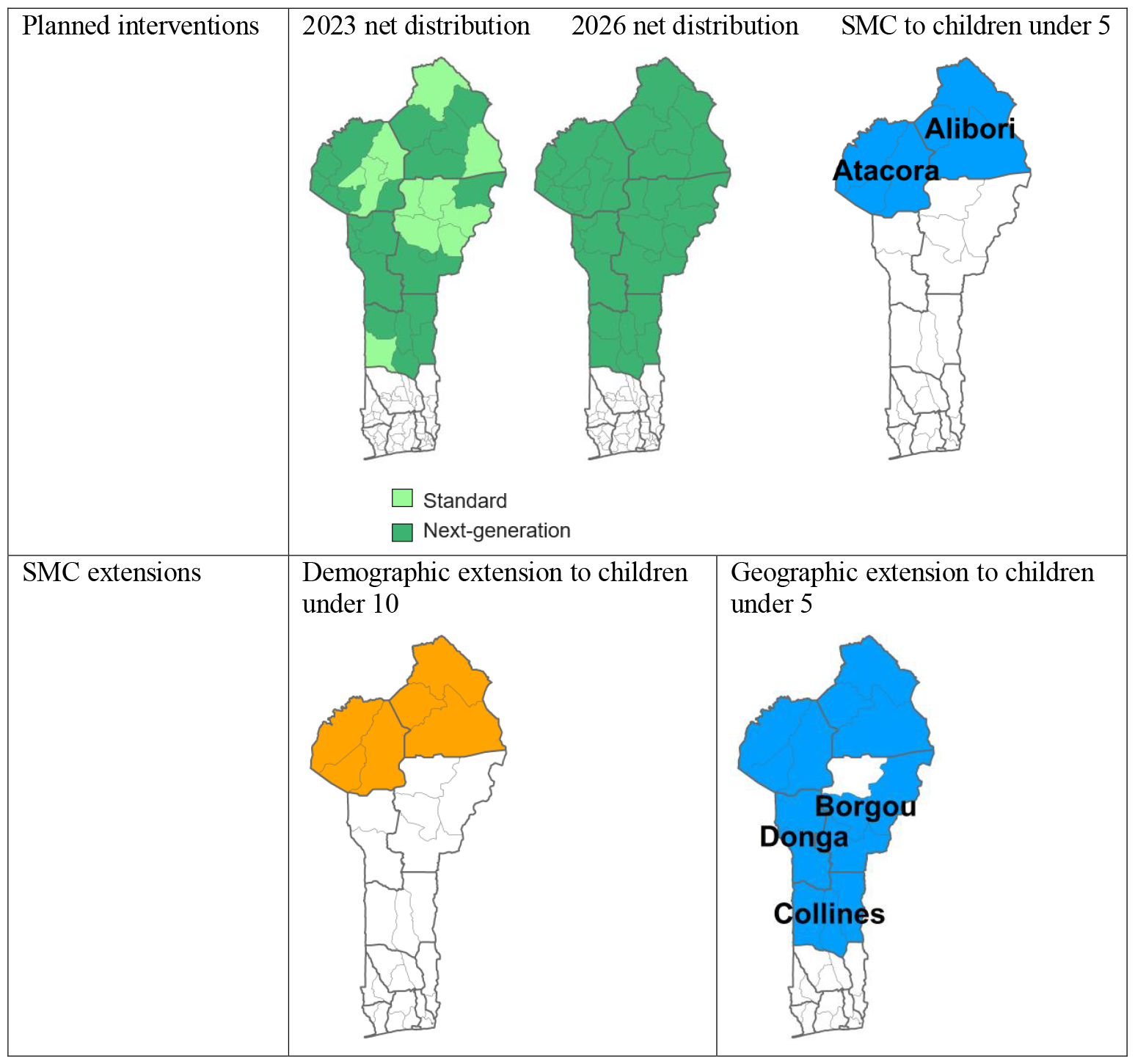
Planned interventions from 2024 and SMC extensions.

| Planned interventions | 2023 net distribution | 2026 net distribution | SMC to children under 5 |
| --- | --- | --- | --- |
| SMC extensions | Demographic extension to children under 10 | Geographic extension to children under 5 |  |

### Comparison of extension scenarios

The indicators of interest were the total number of malaria cases, severe cases, and deaths directly caused by malaria. Since the model accounts for treatment-seeking rates, we obtained both total cases occurring in the community and reported cases at health facilities. We focused on total cases in the community since the treatment seeking behaviour was lower in some departments in 2017 [21], which causes less cases to be reported but does not mean less cases are occurring. We computed cases and deaths averted by each extension scenario by comparing those occurring with only the planned interventions (mass distribution campaigns of ITNs and SMC in children under 5 in Alibori and Atacora) to those with the planned interventions together with extended SMC. We weighted the modelled cases and deaths by the population projected from the 2013 census, with a yearly 3.51% growth rate as measured between 2002 and 2013 [29]. We summed all cases and deaths averted by the demographic extension in Alibori and Atacora between 2024 and 2026, and the cases and deaths averted by the geographic extension in the sanitary zones of Borgou (except Bembereke-Sinende), Collines, and Donga. We also extracted cases averted in each sanitary zone of Borgou, Collines, and Donga for the prioritisation analysis. Mean estimates and credible intervals were obtained from the middle, lower and upper EIR values estimated during the calibration. To account for different population sizes between zones, we also divided the cases and deaths averted by the newly targeted population. For the demographic extension we divided by the number of children between 5 and 10 in Alibori and Atacora, and for the geographic extension by the number of children under 5 in the eligible sanitary zones of Borgou, Collines, and Donga. This way, even if the population growth were stronger than planned, we were comparing cases and deaths averted by additionally targeted population instead of absolute numbers. The number of newly targeted children is also used as a proxy for the extension cost.

## Results

### Greater impact of geographic over demographic extension

The model predicted that 600 (500 - 700) thousand malaria cases in all age groups could be averted between 2024 and 2026 by the demographic extension of SMC (Figure 2). Among these cases averted 3 (−1 - 7) thousand would be severe cases. Additionally, 26 (−9 - 58) direct malaria deaths in all age groups could be averted. By contrast, 1.5 (1.4 −1.5) million cases could be averted by the geographic extension between 2024 and 2026, among these 27 (21 - 33) thousand severe cases. 234 (180 - 279) malaria deaths would also be averted. Between 2024 and 2026 we estimated there would be one million children between 5 and 10 years old in Alibori and Atacora to whom SMC would be additionally administered. In eligible zones of Borgou, Collines, and Donga, 1.8 million children under 5 would be eligible to SMC. When dividing cases averted by the additionally targeted population, we obtained that the demographic extension would avert 609 (530 - 711) malaria cases in all age groups per 1000 targeted children from 5 to 10, and among these 3 (−1 - 7) severe cases (Figure 3). The demographic extension would also avert 25 (−9 - 57) malaria deaths in all age groups per million additionally targeted children. The geographic extension would avert 813 (765 - 845) cases in all age groups per 1000 targeted children under 5, among these 15 (12 - 18) severe cases per 1000 targeted children, as well as 131 (101 - 156) malaria deaths per million targeted children. The geographic extension would thus avert on average 1.33 times more cases, 4.5 times more severe cases, and 5.2 more malaria deaths per targeted child than the demographic extension. Since SMC is recommended to “children belonging to age groups at high risk of severe malaria” [4], the demographic extension would only be more cost-effective than the geographic extension if it were 4.5 times less expensive, or even 5.2 times less expensive when using malaria deaths as the main indicator.

**Figure 2:**
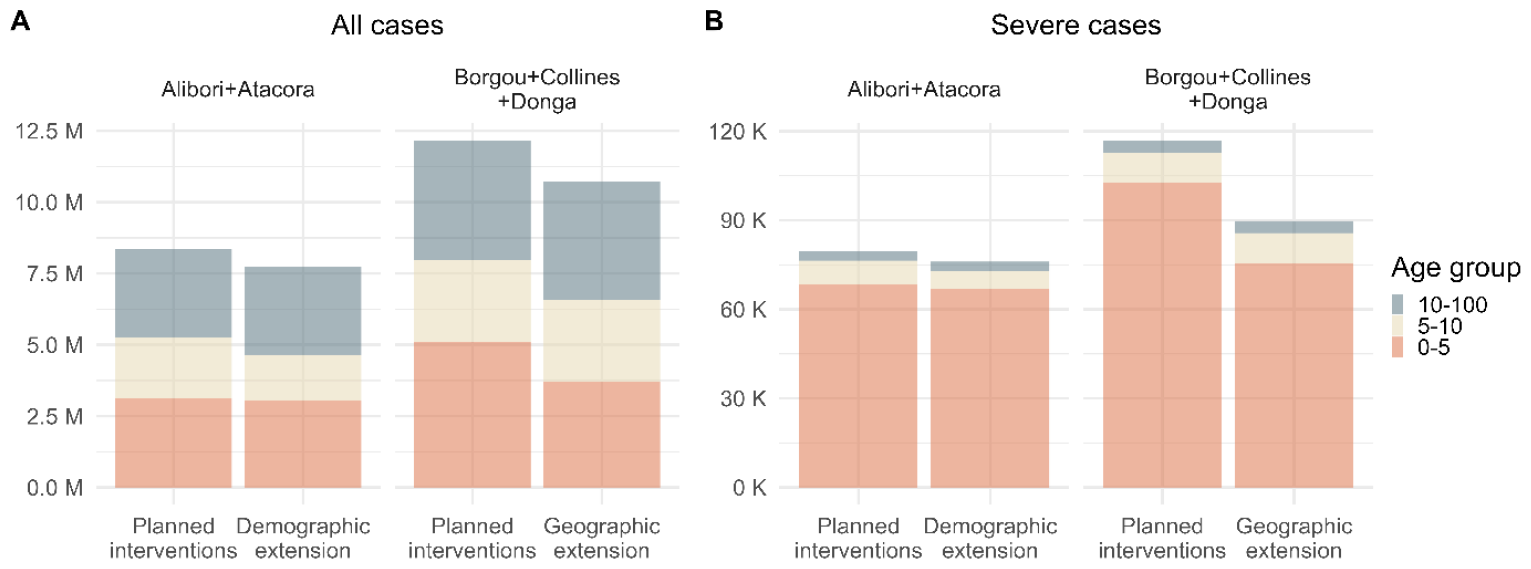
Cases by scenarios and age groups. Distribution of mean predicted malaria cases (A) and severe cases (B) across age groups between 2024 and 2026 with planned interventions and adding SMC extension in eligible zones

**Figure 3:**
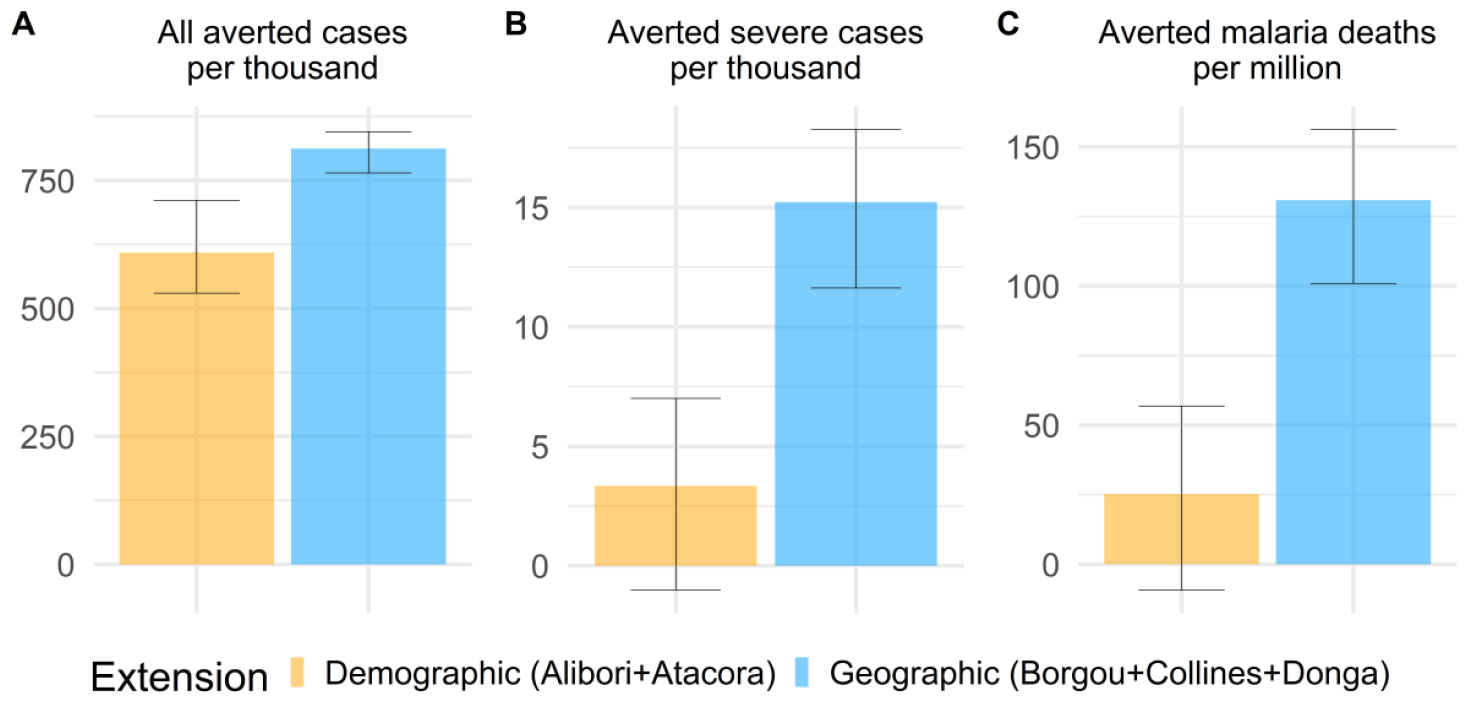
Cases averted per additionally targeted children. A: All malaria cases in all age groups averted by each SMC extension in eligible zones per thousand additionally targeted children (from 5 to 10 for demographic extension and under 5 for geographic extension), between 2024 and 2026; B: Severe cases averted per thousand additionally targeted children; C: Malaria deaths averted per million additionally targeted children. Error bars represent uncertainty on intensity of transmission as well as model stochasticity.

### Prioritisation of sanitary zones among geographic extension

When looking at the number of cases and severe cases averted by the geographic extension of SMC per targeted child by department, no ranking emerged: in Borgou 818 (767 - 852) cases in all age groups could be averted per 1000 children under 5 between 2024 and 2026, among those 16 (12 - 19) severe cases per 1000 children as well as 135 (103 - 163) malaria deaths per million children. In Donga 801 (763 - 826) cases and 14 (11 - 17) severe cases could be averted per 1000 children, as well as 122 (95 - 145) malaria deaths per million children. This similarity was also observed at the operational level of sanitary zones (Figure 4). This means the risks in the different departments and zones are similar enough - or the data are not precise enough to detect any differences - that the decision on where to extend SMC first could be based on operational considerations, such as differences in cost due to the size of the area to cover, or the presence of trained community health workers.

**Figure 4:**
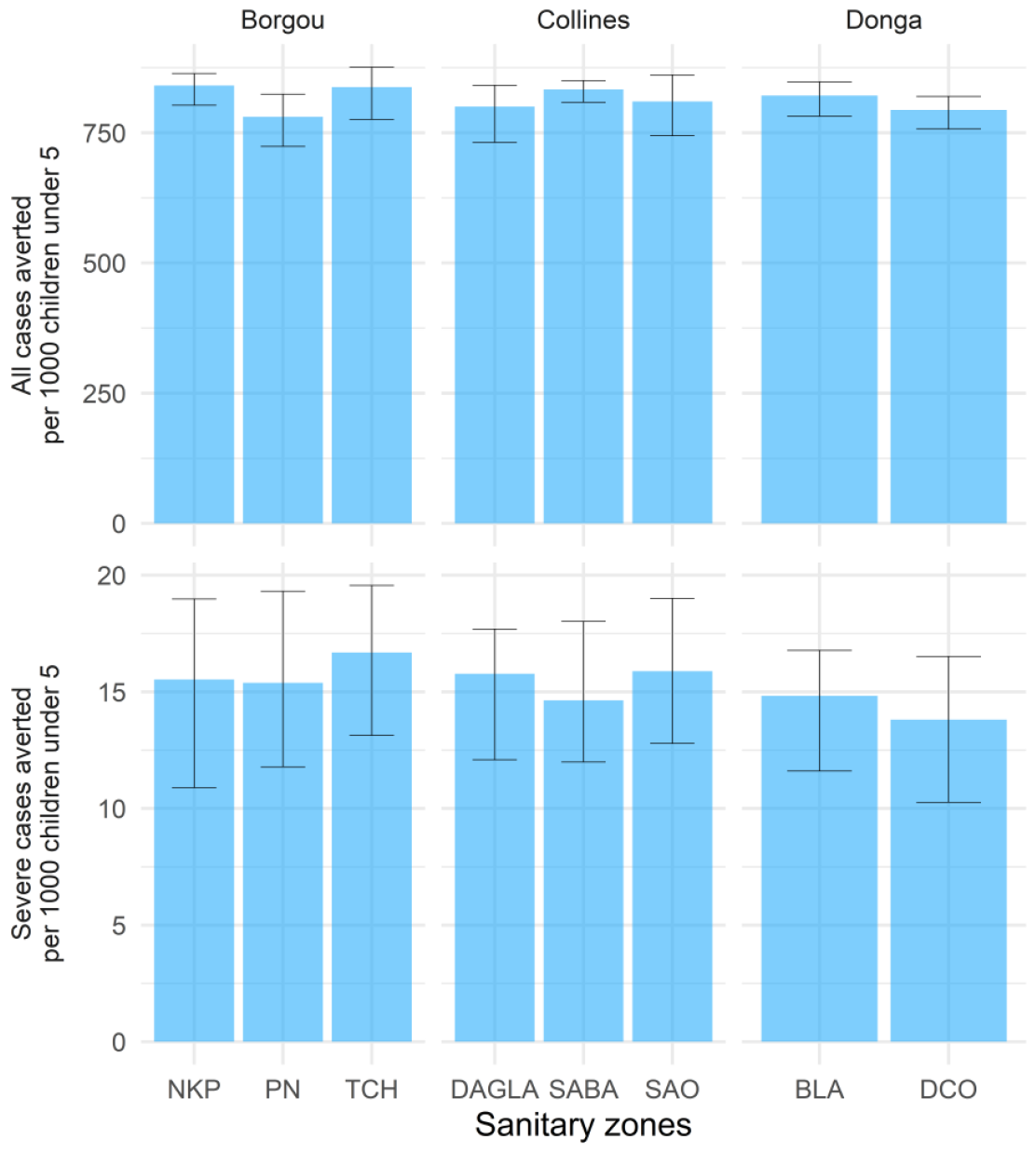
All malaria cases and severe malaria cases averted by the geographical extension of SMC per 1000 children under 5 by sanitary zone. Error bars represent uncertainty on intensity of transmission as well as model stochasticity.

## Discussion

Our results indicate the geographic extension would avert at least four times more severe cases and five times more direct malaria deaths per targeted child compared to the demographic extension. This is consistent with the malaria burden distribution in high-endemic settings where children under 5 are most at risk for malaria, with most malaria severe cases and deaths occurring in children under 5 [11, 12]. However cases averted by target population appear to be similar within the different zones eligible for the geographic extension.

These results were communicated to the NMCP together with a dashboard to explore different epidemiological indicators, and the NMCP decided to include the geographic extension of SMC in the strategic plan. SMC will be administered by community health workers from 2024, and so will first be extended to zones where community health workers have already been recruited and trained, in Donga and Collines. Our results are relevant for and specific to Benin in 2024, but a similar approach could be used to support other countries identifying the most cost-effective implementation of an intervention.

Mathematical modelling can synthetize available evidence on intervention efficacy and extrapolate to specific contexts to support local decision-making. Modelling can account for multiple data sources (seasonality, burden, planned interventions, coverage) to determine which strategy is likely to save most lives. Previous works using mathematical modelling for strategic planning focused on showing the likely impact of already determined combinations of interventions to advocate for funding [14, 30]. This analysis shows that country-specific models can also be used to guide the development of strategies by answering to specific questions of interest for decision makers.

The choice between the two extension scenarios is also influenced by their anticipated relative cost. In 2024 SMC will be administered in Benin by community health workers for the first time, so unit costs from past implementations as campaigns are obsolete. From the cost drivers identified by Pitt et al.[31], only supply chain and sensitisation would likely cost more in the geographic extension since implementation would happen for the first time, but these represent only 1.5% of the total costs and would only be more expensive during the first year of implementation. Purchase of drugs, however, is the second contributor to costs (27.7%) and from six years of age children need to receive one and a half tablet of both SP and AQ, compared to one tablet from 2 to 5 years included [9]. Thus, even without a detailed costing, it is highly likely the unit costs of the demographic extension will be higher than those of the geographic extension. The geographic extension would then be more cost-effective than the demographic extension, especially when considering severe cases and deaths averted. Since cases and severe cases averted per targeted child were similar in zones within the geographic extension, other considerations such as the cost of extending to each zone could have been taken in to account to prioritise where to implement SMC first. In the end the availability of community health workers guided the order of implementation.

This work has a number of limitations. It relies on a model of malaria transmission, which is a simplification of reality. Many parameters of the model were tailored to match local data, which implies that collected data was trusted to reflect reality. EIR was estimated only from prevalence data, and we assumed that all changes in malaria burden were due to interventions, when in practice climate or socio-economics changes also play a role. Given the lack of granularity in the data, some uniformity assumptions were made: net usage and treatment seeking were assumed uniform across each department, *Anopheles* species composition and apparition of resistance identical across the entire country. There are also fixed model parameters, derived from non-Benin specific data: efficacy of nets and treatments, immunity development mechanisms, and case importation. In the model severe malaria episodes can result from high parasitaemia and/or co-infection, the risk of the latter increasing with EIR and decreasing with age [27]. Since in northern Benin the risk of malaria is high, we estimated high EIR values and thus the model attributed most severe malaria episodes to co-infections, which occur mostly in young children. This explains why our results attribute around 90% of severe cases to children under 5.

We reproduced the efficacy of SMC observed in a randomised controlled trial [28], though in real-life settings SMC will likely avert less cases due to less controlled implementation. However, this challenge will probably affect the implementation of geographic and demographic extensions in a similar manner so correcting for this simplification is unlikely to change our results. We assumed SP-AQ had the same efficacy in children under and above 5 years of age, for lack of age-specific data on efficacy. If the drug efficacy varies with age this will in turn affect our results in the same direction (more cases averted in the age groups with higher efficacy). However similar odds ratios of mortality and incidence during transmission seasons [5] seem to indicate efficacy is similar in both age groups. Moreover the difference in severe cases averted is so strong that the efficacy in older children would have to be at least four times higher to change our conclusions, which is unlikely. It is crucial for mathematical models to be able to reproduce the effect sizes of SMC in different ages to guide decisions.

Geographic heterogeneity in future SMC coverage and population structure could not be included for lack of data. We assumed SMC coverage would stay constant through time, space, and age groups. In Senegal a higher coverage in older children was observed by a few percentage points [9], but even if these results were valid for Benin the difference would not be strong enough to change our conclusions.

Population estimates rely on the 2013 census [29], with a 3.51% national growth rate applied to each commune, which was the method recommended by the NMCP. These estimates are outdated: a population count before the 2020 mass net distribution campaign found the population to be 13.6% larger than projected [32], but at the time of analysis results from the new census had not been published. Population growth is also not uniform across departments: between 2002 and 2013, population yearly increased by 2.5% in Collines and 4.6% in Alibori. Since we divided cases averted by the target population, these issues should not influence our results. The proportions of children under 5 (17.4%) and between 5 and 10 (13.7%) are also taken from the 2013 census. They appear not to have changed drastically during the past decade [33].

In Benin, the geographic extension of SMC to southern departments in 2024 would avert at least four times more severe cases per additionally targeted child than its demographic extension, and five times more direct malaria deaths. In zones eligible to the geographic extension SMC was predicted to avert similar number of cases and severe cases per child under 5. The NMCP adopted the recommendations in their strategy and secured funding for the geographic extension starting in 2024. This country-led collaboration, together with a country-specific modelling approach, led to evidence-based strategic decisions which could be fully funded.

## Data Availability

Input data for simulations, simulation datasets and additional data necessary for analysis such as population are available upon reasonable request to the authors.

## Acknowledgments

The authors thank Tatiana Alonso Amor, Mar Velarde, Flavia Camponovo, Monica Golumbeanu, Didier Adjakidje for valuable discussions, George Shireff for past modelling analyses in Benin, Thomas Smith for proofreading the manuscript. Calculations were performed at sciCORE (http://scicore.unibas.ch/) scientific computing centre at the University of Basel (Basel, Switzerland).

