## Supplementary material for "Modelling to support decisions about the geographic and demographic extensions of seasonal malaria chemoprevention in Benin"

### A. Calibration workflow

Country data

Seasonality

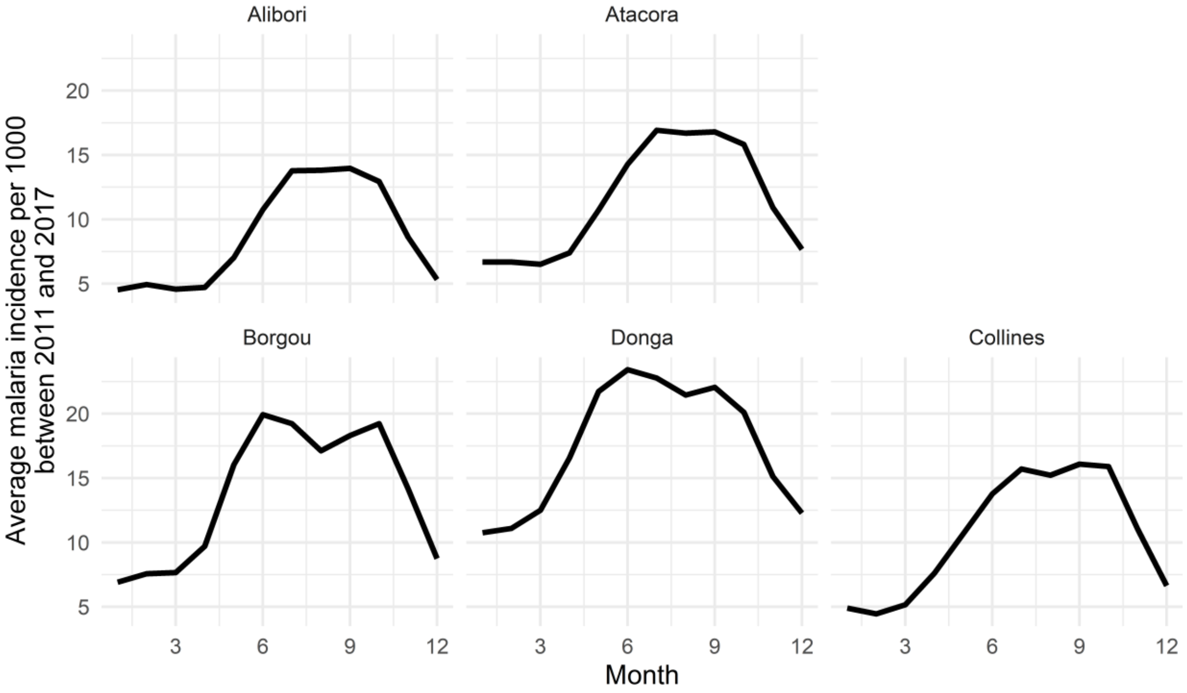

**Figure A.1: Monthly incidence by department averaged between 2011 and 2017.**
Reported incidence was calculated using monthly DHIS2 confirmed cases averaged between 2012 and 2017 and population estimates from 2013 census.[1]

Insecticide-Treated Nets

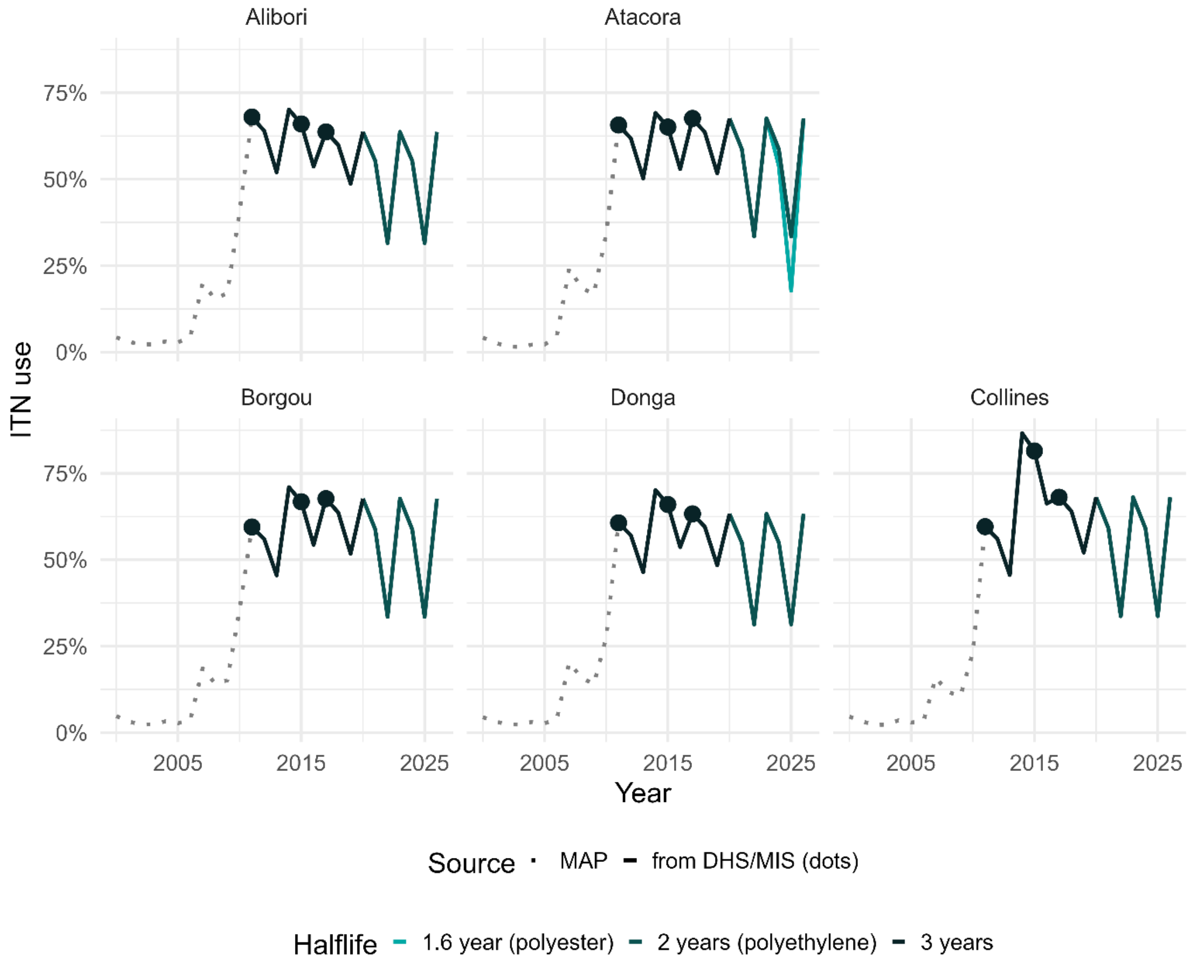

**Figure A.2: ITN use assumptions by department.**
Until 2011 we use the yearly estimates from the Malaria Atlas Project, from 2011 to 2019 we use the DHS and MIS surveys, and assume nets have a three-year halflife in-between distributions. From 2020 we kept the initial use as measured after the 2017 campaign but adjusted the halflife to the fabric of nets distributed. In 2023 two types of nets (polyester and polyethylene) were distributed in Atacora, and we modelled the correct type in each commune of the department.

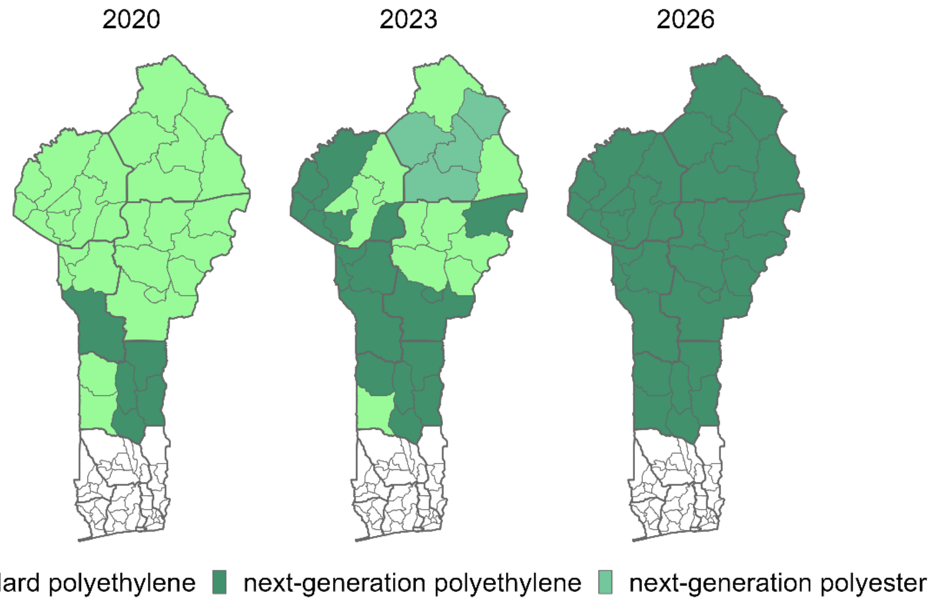

**Figure A.3: Type of nets distributed or planned to be distributed in 2020, 2023 and 2026**

IRS

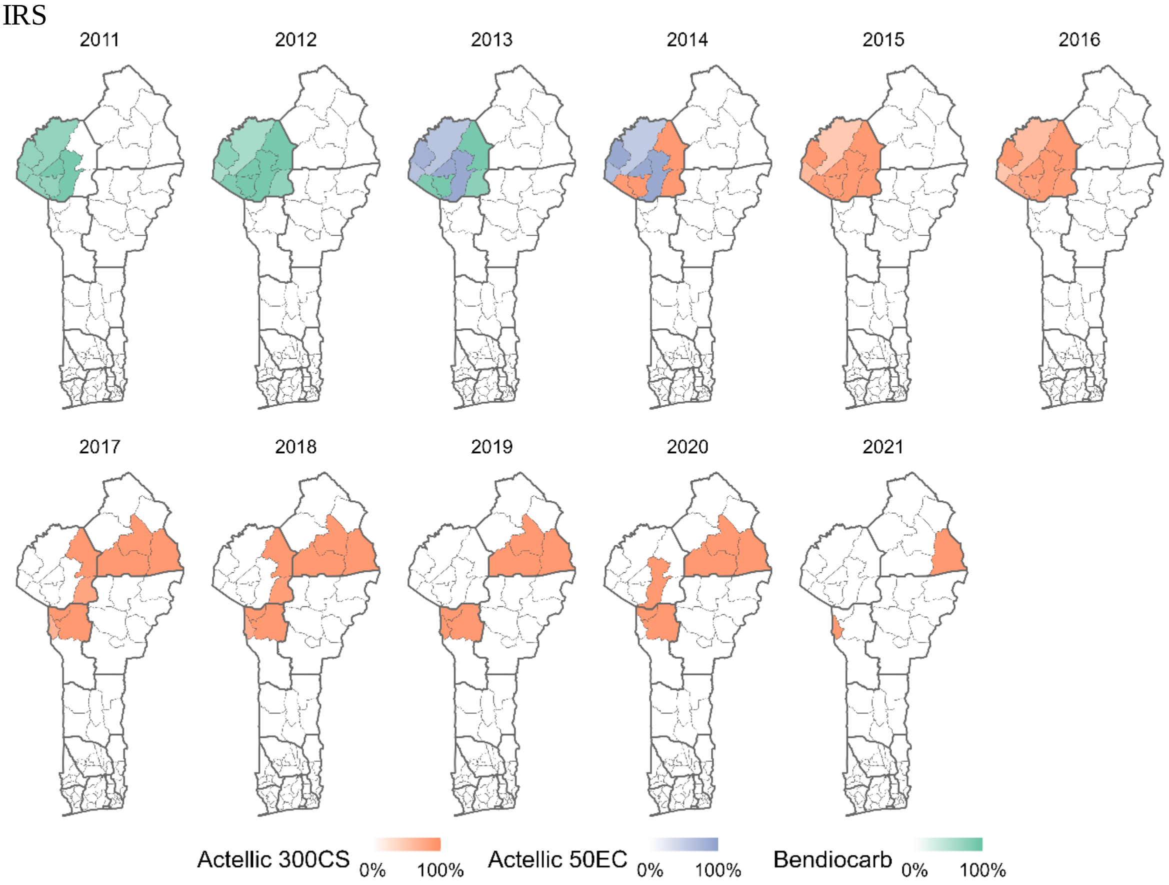

**Figure A.4: Assumptions for ingredients and population coverage of IRS campaigns between 2011 and 2021.**

In 2020 and 2021 Pirimiphos-methyl was sprayed but the most recent parameterisation available was for Actellic 300CS.

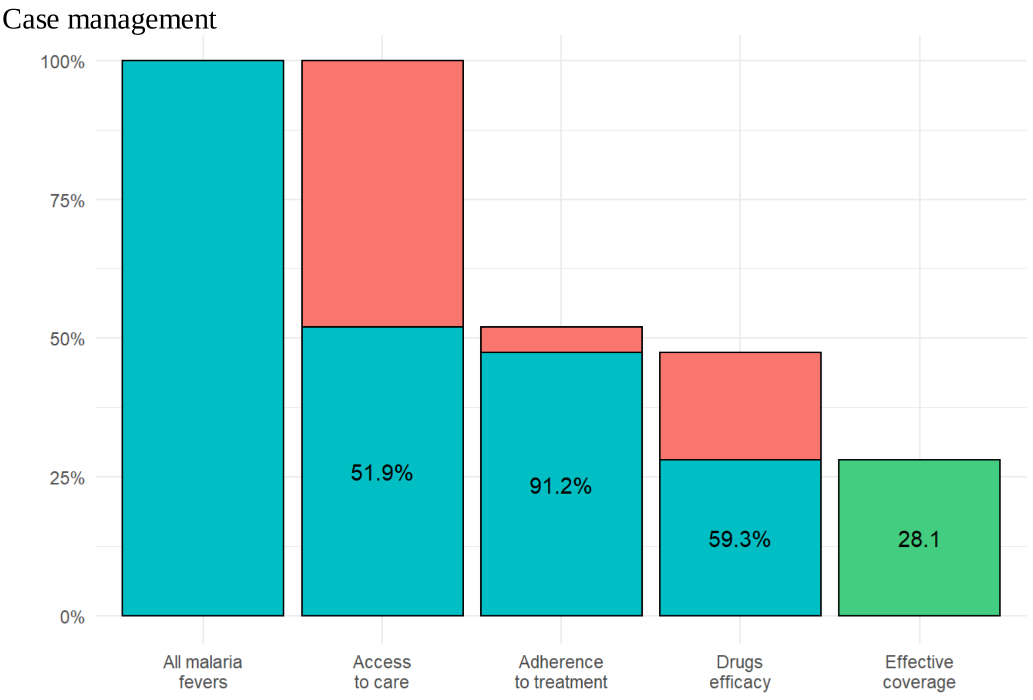

**Figure A.5: Case management cascade from access to care measured in 2017-2018 DHS[2] at the national level.**

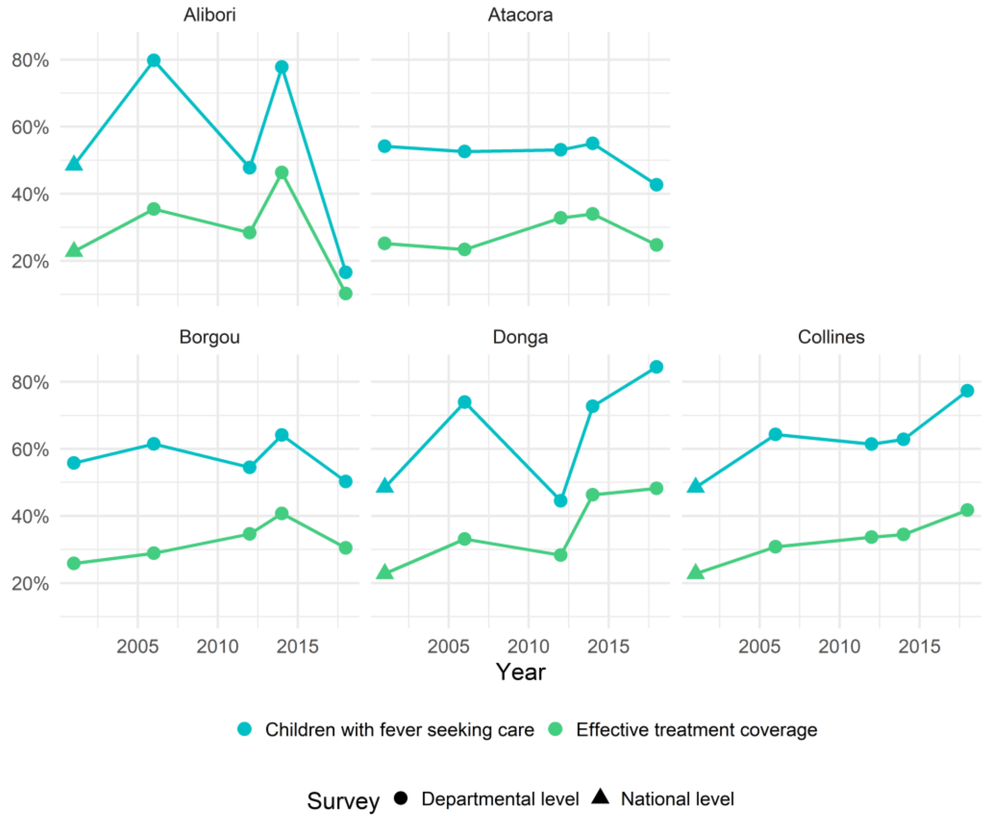

**Figure A.6: Evolution of access to care (blue) and effective treatment coverage (green) in the five departments of interest.**

SMC

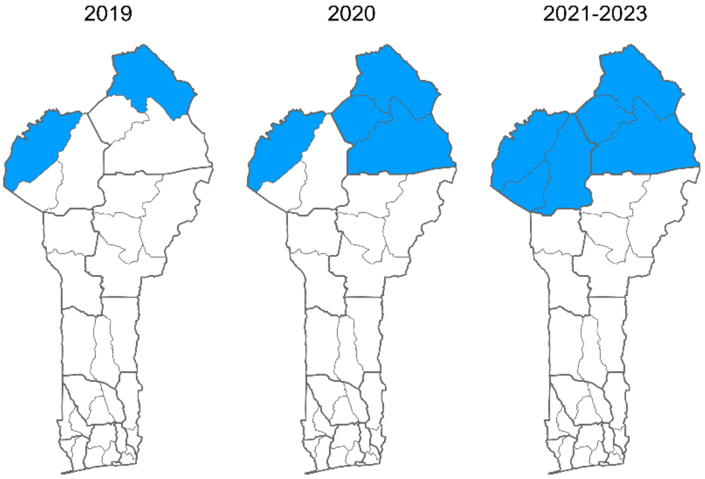

**Figure A.7: SMC implementation in children under 5 between 2019 and 2023**

Age structure

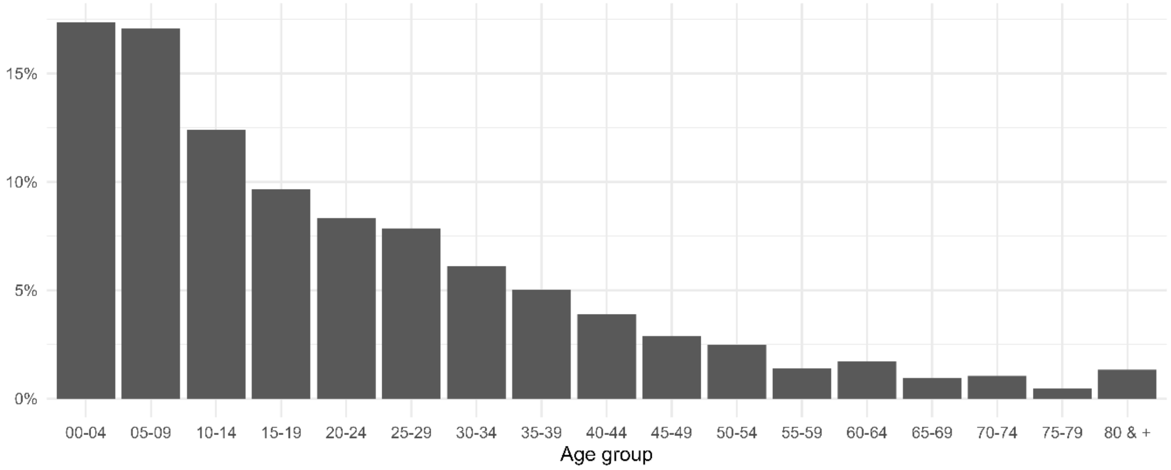

**Figure A.8: Proportion of population in each age group in Benin according to 2013 census**[1]

Simulations

2000-2019

Before 2000, the model is assumed to be at equilibrium. We then include the history of malaria control interventions deployed between 2000 and 2019, namely vector control, case management and SMC. Since household surveys are powered at the department level, history of interventions is simulated at the department level except when information is available at the lower commune level, such as for IRS and SMC.

Parasite detection limit is set at 100 parasites per microliter to reproduce RDT detection limit.[3] Since we only need prevalence for the calibration step and no rarer events such as severe cases or deaths, we simulate a small population of 3000 individuals. This is also a trade-off to avoid running many long simulations.

Calibration

Calibration was done by running a grid of simulations sharing the same history of interventions and geographical characteristics but with EIR values ranging from 1 to 250 (with a step of 1 until 10, and a step of 2 afterward), and for 10 seeds.

We note $X^{obs} = \left(X_i^{obs}\right)_{i\in\{2006,\ldots,2019\}}$ the estimates for prevalence for the years 2006 to 2019 with their associated uncertainty estimate for standard deviation noted $\sigma = (\sigma_i)_{i\in\{2006,\ldots,2019\}}$. The standard deviations were estimated from the 95%-confidence intervals $(X_i^{low}, X_i^{up})$ by $\sigma_i = \frac{\max(X_i^{obs}-X_i^{low}, X_i^{obs}+X_i^{up})}{1.96}$. For a given set of parameters θ (including the initial EIR), the associated OpenMalaria simulation for prevalence over the years 2006 to 2019 is noted $X^{sim,\theta} = \left(X_i^{sim,\theta}\right)_{i\in\{2006,\ldots,2019\}}$. For each simulation in the grid, a pseudo-likelihood is computed to compare the predicted prevalence to the reported prevalence:

$$L\left( X^{obs}, X^{sim,\theta}\right) = \Pi_{i=2006}^{2019} \Phi_{X_i^{sim,\theta}, \sigma_i}\left(X_i^{obs}\right)$$

where $\Phi_{\mu,,\sigma}$ represents the probability density function of a normal distribution with mean μ and standard deviation σ. Because the standard deviation associated with each observation point is accounted for, the pseudo-likelihood assigns more importance to observations with lower uncertainty. The EIR value with highest log-likelihood ($EIR^{MLE}$) is selected as point estimate.

An uncertainty estimate is computed using the methodology by Ionides et al.,[4] adapted to this particular use case. In Ionides et al., a modified profile likelihood approach is used, where a cut-off on the profile likelihood defines a confidence interval on the parameter of interest. The cut-off, based on the quantile of a Chi square distribution, is adjusted to account for uncertainty in the pseudo-likelihood function. For this, a quadratic approximation is fitted on the smoothed likelihood and the parameters of that quadratic function are used to calculate the adjustment factor.

This approach is extrapolated to our context and definition of the pseudo-likelihood. The method is applied for each administrative area using 10 stochastic replications, with a loess smoother equal to 1, but only restricted to the EIR values for which $\frac{|EIR^{MLE}-EIR|}{EIR^{MLE}} \leq c = 0.5$ . This restriction permitted an adaptive window for which the quadratic approximation is valid for all administrative areas simultaneously, thus avoiding having to choose a loess smoother for each administrative area independently. When $EIR^{MLE}$ is above (respectively below) $\lambda = 1$, the highest (resp. lowest) EIR value of the grid is retained as upper (resp. lower) uncertainty bound.

The EIR point estimate as well as the upper and lower uncertainty bounds are all simulated for the future scenarios, in order to propagate the uncertainty in transmission intensity due to prevalence data fitting.

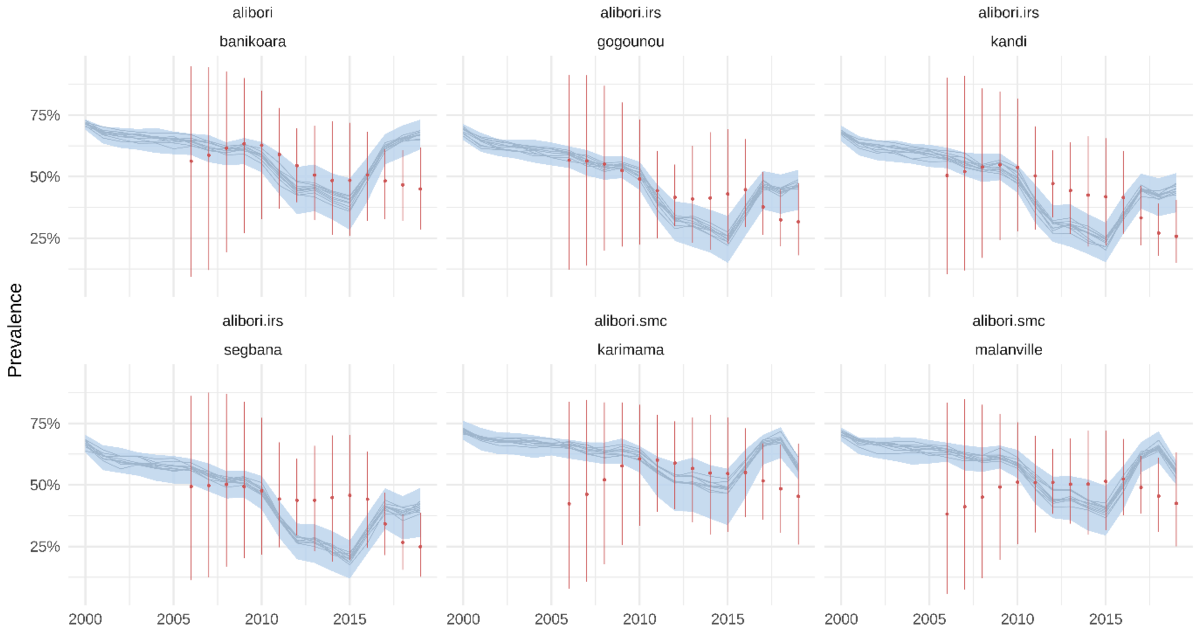

**Figure A.9: Calibrated simulations for communes of Alibori.**

Commune-level prevalence estimates in children from 2 to 10 (in red) and calibrated simulations (in blue) for each commune. Individual simulation runs are represented as well as estimated uncertainty.

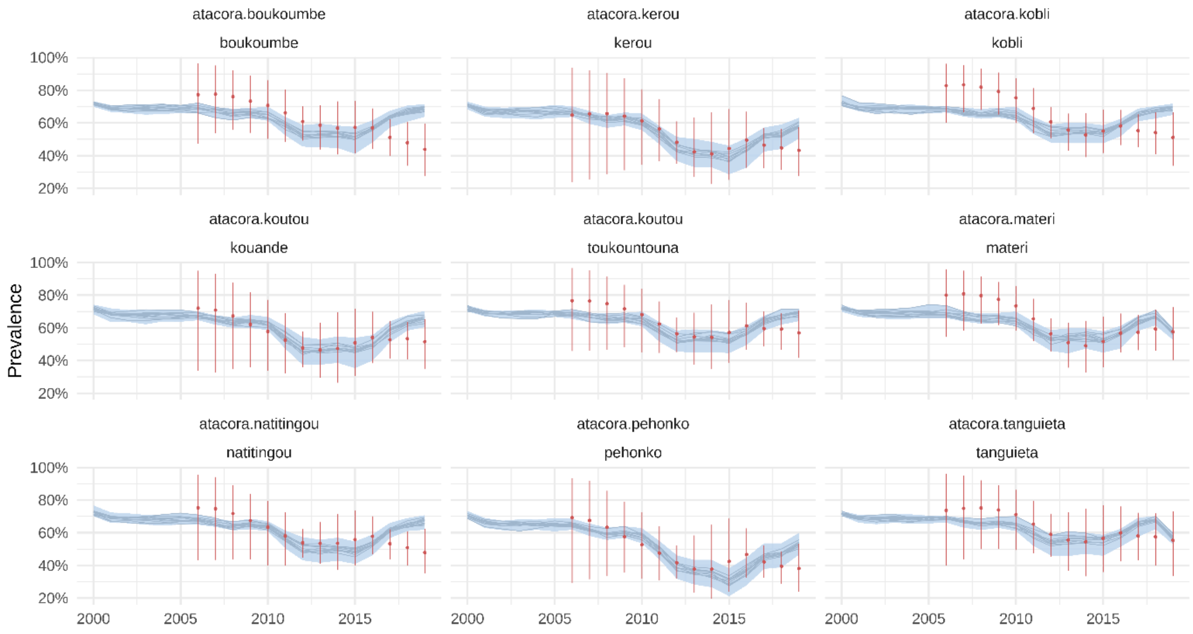

**Figure A.10: Calibrated simulations for communes of Atacora.**
Commune-level prevalence estimates in children from 2 to 10 (in red) and calibrated simulations (in blue) for each commune. Individual simulation runs are represented as well as estimated uncertainty.

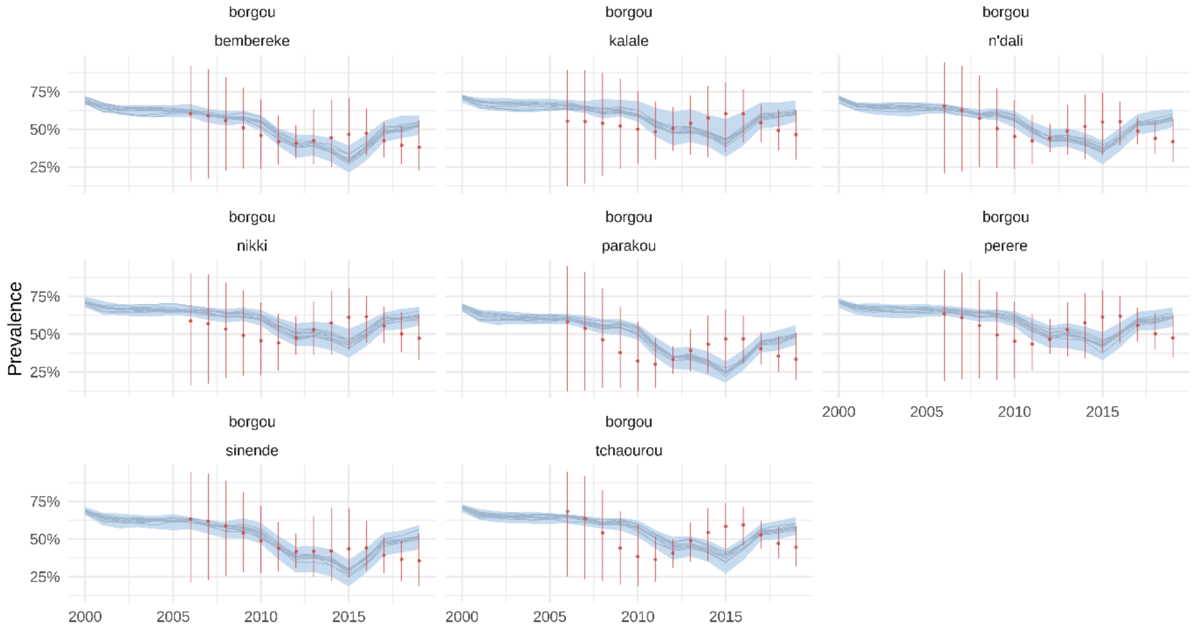

**Figure A.11: Calibrated simulations for communes of Borgou.**
Commune-level prevalence estimates in children from 2 to 10 (in red) and calibrated simulations (in blue) for each commune. Individual simulation runs are represented as well as estimated uncertainty.

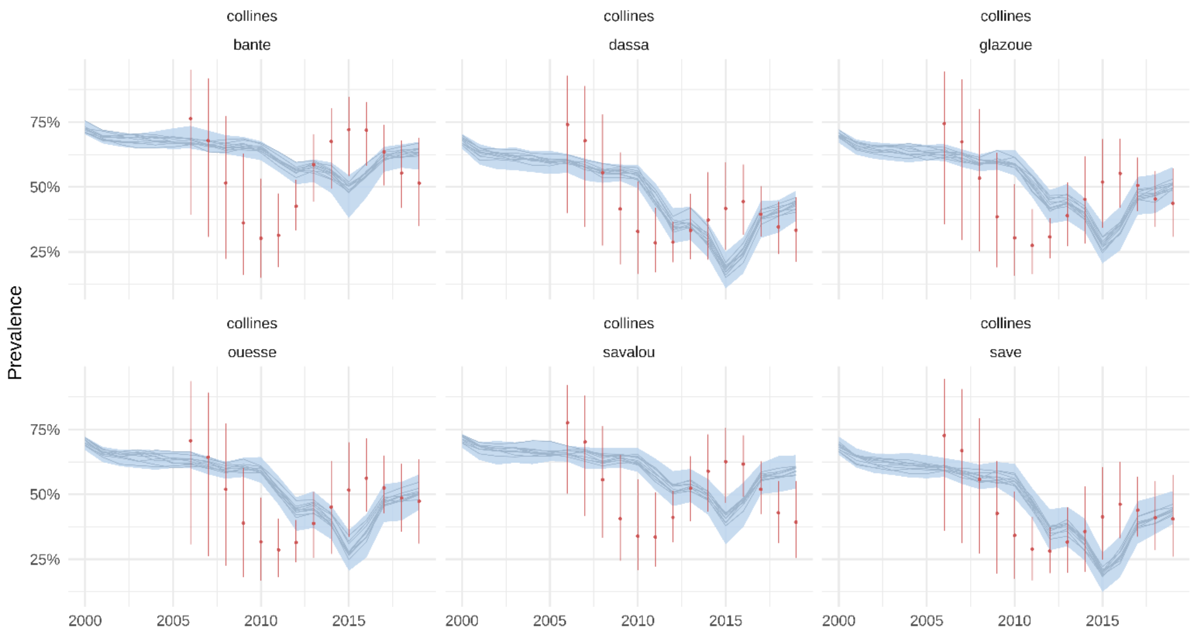

**Figure A.12: Calibrated simulations for communes of Collines.**
Commune-level prevalence estimates in children from 2 to 10 (in red) and calibrated simulations (in blue) for each commune. Individual simulation runs are represented as well as estimated uncertainty.

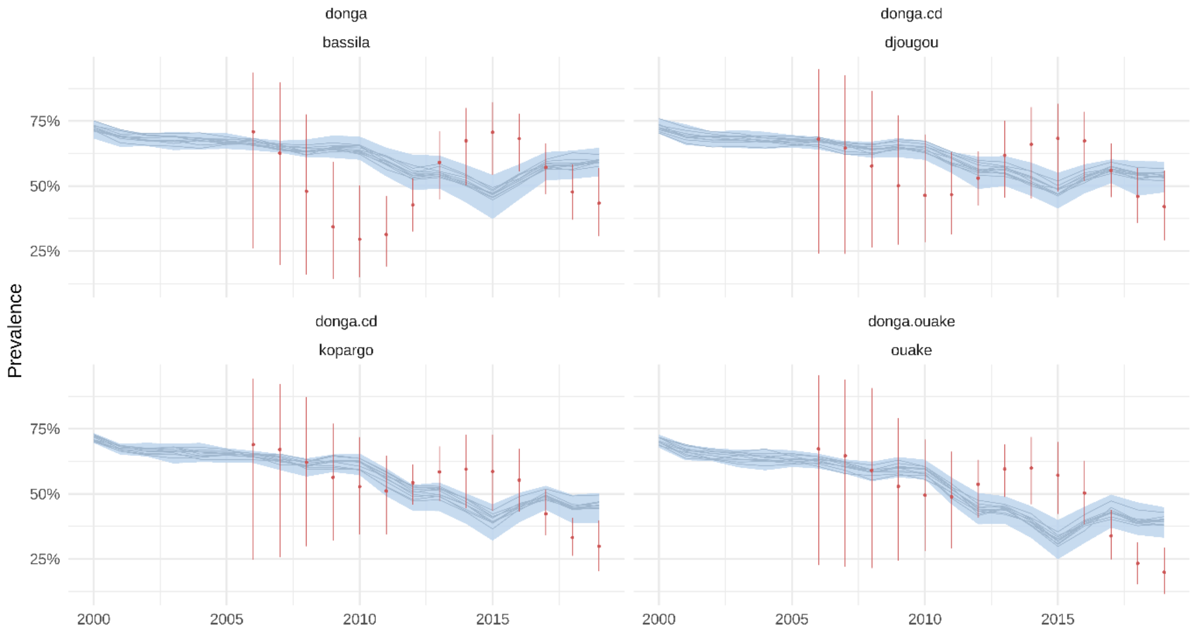

**Figure A.13: Calibrated simulations for communes of Donga.**
Commune-level prevalence estimates in children from 2 to 10 (in red) and calibrated simulations (in blue) for each commune. Individual simulation runs are represented as well as estimated uncertainty.

2020-2023

We simulate interventions from 2020 (either past or planned) with the three EIR values (point estimate, lower and upper bound) per commune estimated during the calibration. Since we now have less simulations we simulate a 50000 population in each commune, which was the population of the least-populated commune in 2020 (Toukountouna) and is enough to reliably estimate severe cases and deaths. Interventions include 2020 and 2023 mass distribution campaigns, SMC and IRS spraying. Case management values remain constant from 2017.

Future scenarios are modelled similarly, starting from 2024.

Validation

Since we used prevalence estimates to fit the transmission intensity, we could validate the model predictions against reported data on cases and deaths. We weighted cases and direct malaria deaths predicted by the model by the population of each commune. The population was estimated from the 2013 census[1] at the commune level, applying a 3·51% annual growth rate to each commune (national growth rate between 2002 and 2013 censuses).

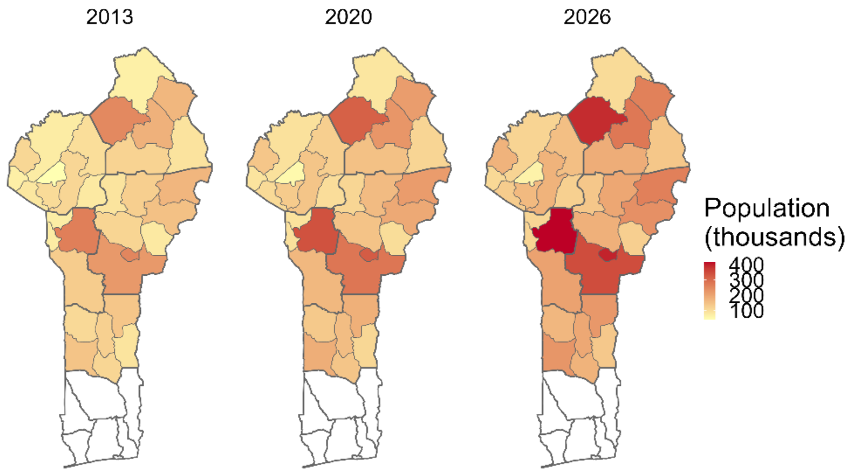

**Figure A.14: Population by commune in 2013, 2020 and 2026**

Modelled cases and deaths were compared to reported incidence from DHIS2 as well as estimated incidence of cases and deaths in the World Malaria Report 2022[5] to make sure the orders of magnitude were in agreement.

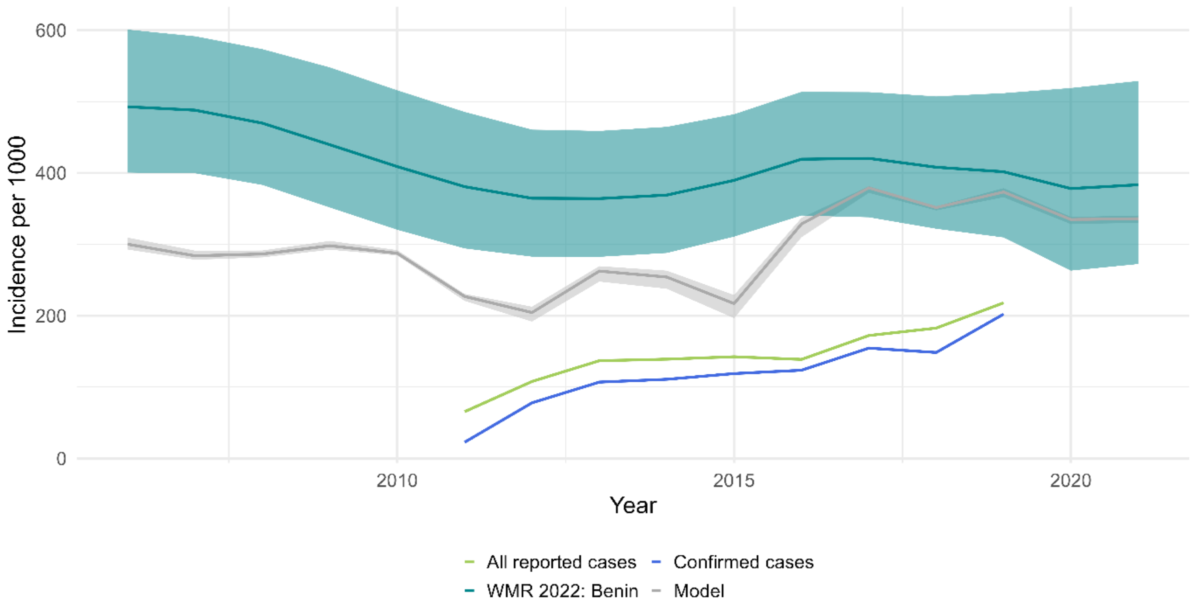

**Figure A.15: Modelled reported incidence against DHIS2 incidence (all reported cases and only confirmed cases) and estimates from World Malaria Report 2022**

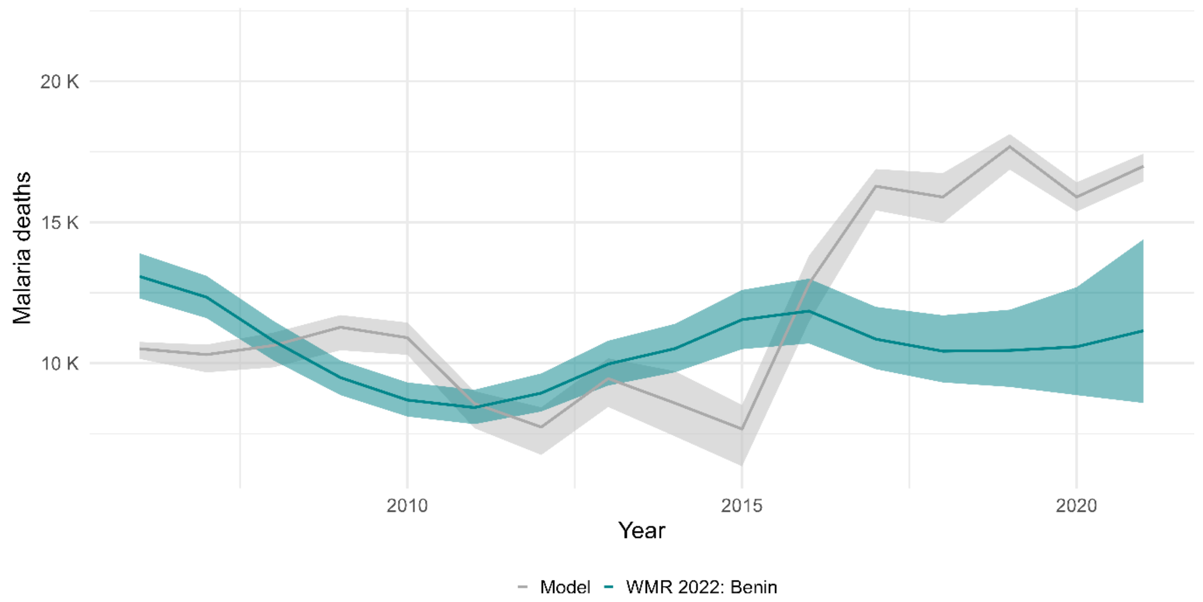

**Figure A.16: Modelled directed malaria deaths against estimates from World Malaria Report 2022**

### B. Additional results

Effect sizes

We compare the reduction in cases and severe cases induced by both SMC extension scenarios compared to planned interventions only.

In Alibori and Atacora the model predicted 2·1 (2·0 - 2·3) million cases in children from 5 to 10 years old between 2024 and 2026 with the scheduled interventions, and 1·6 (1·5 - 1·7) with the demographic extension of SMC (see Figure 2 in main paper). This would represent a 25·4% (24·1 - 27) reduction in cases and 24·4% (19·8 - 30·0) in severe cases in children aged 5 to 10.

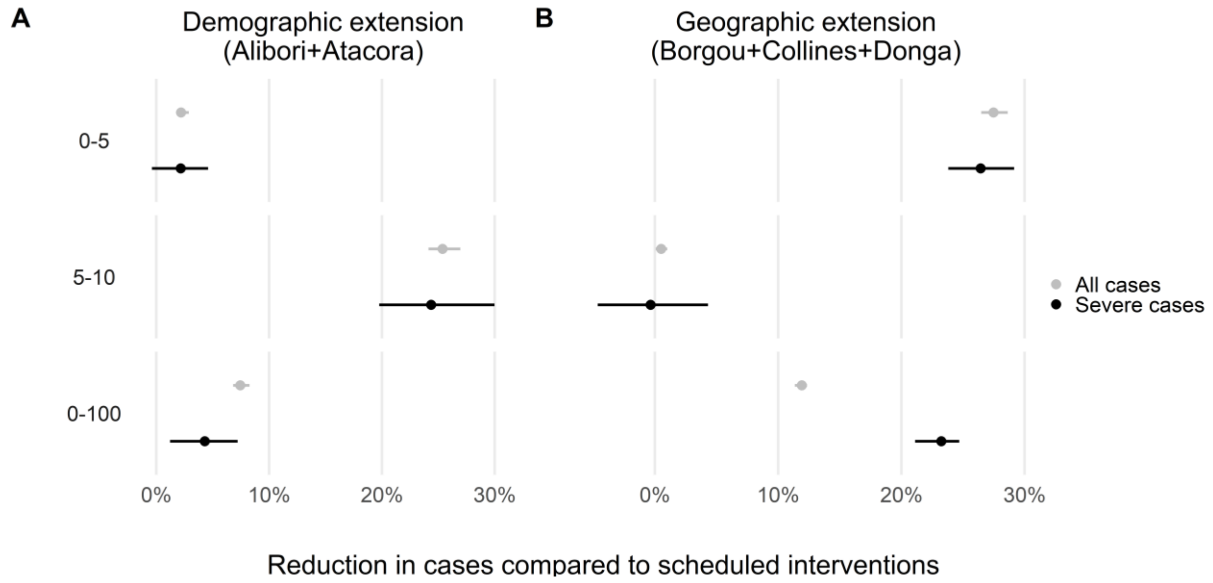

**Figure B.1: Percentage of reduction of all predicted malaria cases or severe cases induced by SMC demographic (A) or geographic (B) extension between 2024 and 2026 by age group.** Error bars represent uncertainty on intensity of transmission as well as model stochasticity.

In zones eligible to the geographic extension, the model predicted 5·1 (4·7 - 5·4) million cases in children under 5 between 2024 and 2026 under the scheduled interventions, against 3·7 (3·4 - 4) million with SMC. This represents a 27·5% (26·5 - 28·6) reduction in cases under 5 due to SMC geographic extension, and a 26·4% (23·8 - 29·2) reduction in severe cases when comparing scheduled interventions to SMC geographic extension.

The impact of the demographic extension on the general population would be smaller than the one of the geographical extension, with cases in all ages diminishing by 7·4% (6·8 - 8·3) and severe cases by 4·3% (1·2 -7·2) with the geographic extension compared to scheduled interventions, respectively 11·9% (11·4 - 12·4) and 23·2% (21·1 - 24·7) with the geographic extension.

Averted cases

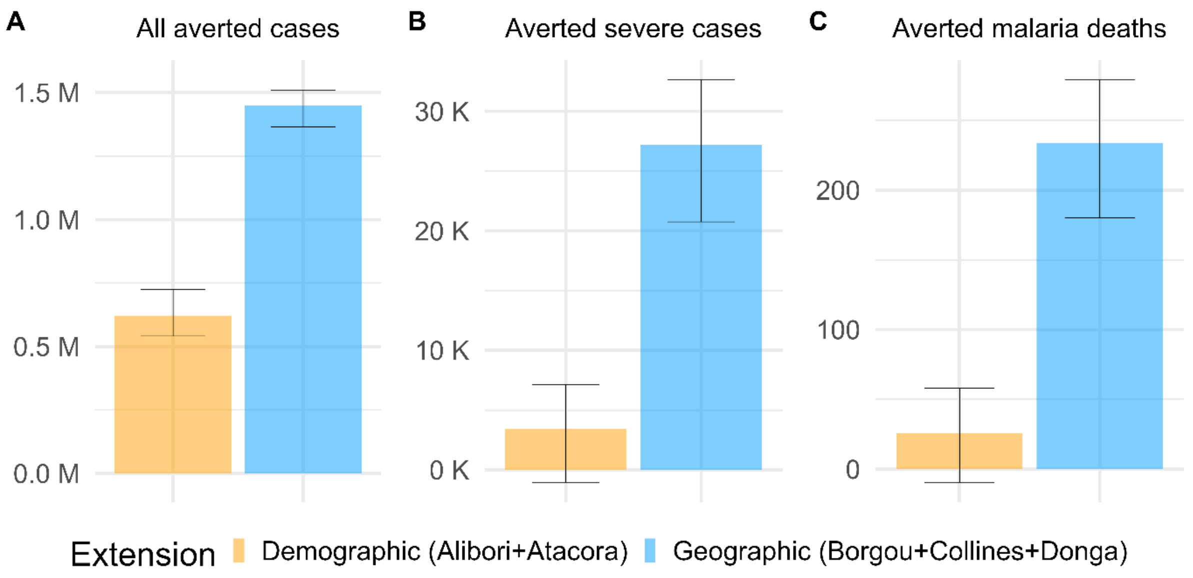

**Figure B.2: Absolute averted cases, severe cases and deaths by each extension scenario.**
A: All cases in all age groups averted by each SMC extension in eligible zones between 2024 and 2026; B: Severe cases averted between 2024 and 2026; C: Malaria deaths averted between 2024 and 2026. Error bars represent uncertainty on intensity of transmission as well as model stochasticity.
